# Harnessing exhaled breath for lung cancer early detection – results from the ExPeL study

**DOI:** 10.64898/2026.03.19.26348785

**Authors:** Danita Patel, Leon D’Cruz, Waqar Ahmed, Anoop Chauhan, Nawar Bakerly, Seamus Grundy, Drupad Trivedi, Sean Knight

## Abstract

**Background:** Scalable, non-invasive tools are critically needed to improve early lung cancer detection and optimize primary care referral pathways. We evaluated Inflammacheck®, a point-of-care device utilizing exhaled breath condensate (EBC) H₂O₂ and physiological parameters with machine learning, for non-invasive lung cancer detection in a real-world screening population.

**Methods:** ExPeL study participants, from the UK Targeted Lung Health Check (TLHC) programme, included individuals with suspected lung cancer and low-risk ever-smoker controls. EBC was collected via Inflammacheck®, measuring H₂O₂, end-tidal CO₂, humidity, temperature, and exhalation flow rate. Multivariate analyses (PCA, LDA, Mahalanobis distance) assessed intrinsic group separation. SMOTE-balanced data trained supervised machine learning models (stacked and voting ensembles), which were then evaluated on held-out test sets. In parallel, untargeted LC–MS metabolomics was performed to identify discriminatory molecular features.

**Results:** Analysing 34 participants with valid EBC data, 83% of cancer cases were early-stage (I–II), reflecting a screening population. Multivariate analysis clearly separated lung cancer and controls across PCA, LDA, and Mahalanobis mapping. The voting ensemble model achieved: Accuracy 85.7%, Sensitivity 80%, Specificity 100%, Precision (PPV) 100%, ROC–AUC 0.90, MCC 0.73. Crucially, no false positives were identified. EBC variables revealed greater dispersion in cancer patients, reflecting physiological heterogeneity missed by univariate analysis. Untargeted metabolomics identified 2,132 features, with four key metabolites yielding an AUC of 0.969 for cancer discrimination.

**Discussion:** Inflammacheck® effectively distinguishes early-stage lung cancer via a rapid, non-invasive breath test, findings which are highly relevant for primary care and screening triage, where non-specific symptoms and low prevalence pose challenges.

## INTRODUCTION

For cost efficiency, most lung cancer screening programs use a pre-screen clinical score to enrich a high-risk population. In the UK, this means that only 40% of people with lung cancer would have been eligible for CT screening prior to their diagnosis (1). New tests are needed to focus provision of CT scans more effectively. Expired air contains a rich source of molecules that can be sampled non-invasively to discover new tests. Rapid cooling of breath leads to exhaled breath condensate (EBC), which contains sediments of aerosolised particles. EBC analysis has previously identified metabolic compounds associated with late-stage lung cancer (2), but it is not clear whether these extrapolate to early stage disease (3). Reactive oxygen species (ROS) are promising biomarkers as they are readily produced in the tumour microenvironment by both cancer and immune cells (4). Hydrogen peroxide in EBC was significantly elevated in patients with non-small cell lung cancer compared to controls in two studies (5,6). However, both studies excluded patients with chronic respiratory disorders, which independently increase ROS in EBC as reviewed previously (7).

Inflammacheck® is a handheld device that measures hydrogen peroxide via an enzymatic reaction within a sensor, providing point-of-care, accurate measurements. It also measures breath parameters in tidal breathing. This constellation of readings was exploited in the VICTORY study to develop a machine learning algorithm capable of distinguishing lung cancer from chronic respiratory disorders (8).

In ExPeL we extend the findings from VICTORY, testing the diagnostic accuracy of Inflammacheck® in populations relevant for lung cancer screening. In parallel, we have performed an unbiased screen of EBC for novel molecular candidates, identifying new metabolites associated with lung cancer that will be incorporated into Inflammacheck® in the future.

## METHODS

### Study Design and Participant Recruitment

The Harnessing Exhaled Hydrogen Peroxide for Early Lung Cancer Detection (ExPeL) study received ethical approval from the West Midlands NHS Research Ethics Committee (IRAS: 336691; REC: 24/WM/0028; ISRCTN: 81020233). All participants provided written informed consent.

Participants aged ≥16 years were recruited from the Greater Manchester Lung Cancer Screening service. **Cases** comprised individuals with radiologically suspected lung cancer referred for CT-guided biopsy following computed tomography (CT) imaging undertaken within the Targeted Lung Health Check (TLHC) programme, for respiratory or cancer-related symptoms (“symptomatic”), or for non-respiratory indications (“incidental”). TLHC is part of NHS England’s targeted screening initiative for ever-smokers, designed to improve early lung cancer detection in high-risk populations (9).

**Controls** were defined as ever-smokers assessed through the TLHC pathway who underwent low-dose CT imaging and were not diagnosed with lung cancer following guideline-directed radiological assessment and follow-up. This included individuals with pulmonary nodules classified as low risk according to national nodule management criteria, and one participant with histologically confirmed benign disease. We reiterate that the control group represents screen-negative, low-risk TLHC participants, not lifelong benign controls. Detailed justification of control classification is provided in the online-Supplementary file.

### Exhaled Breath Condensate Collection

Exhaled breath condensate (EBC) was collected using the Inflammacheck® device. Participants breathed tidally through a disposable mouthpiece while wearing a nose clip for up to six minutes; sampling terminated automatically once sufficient condensate was obtained. Five parameters were recorded: hydrogen peroxide concentration (µM), peak end-tidal carbon dioxide (%), peak humidity (%), peak breath temperature (°C), and mean exhalation flow rate (L/min).

A subset of participants also provided an additional EBC sample using the Coronacheck® device to enable larger-volume collection for downstream metabolomic analysis. EBC measurements were obtained for research purposes only and did not inform clinical management.

### Data Quality Control and Sample Exclusion

Following quality review, a sensor equilibration issue affecting a subset of Inflammacheck® devices was identified, resulting in unreliable humidity-dependent measurements. The protocol was amended and additional participants recruited. Of 56 participants undergoing Inflammacheck® testing, 15 datasets were excluded due to sensor artefact and 7 due to insufficient sample acquisition, leaving 34 valid datasets for analysis.

### Multivariate (PCA, LDA and Mahalanobis distance analysis) Analysis

All EBC variables were standardised to zero mean and unit variance prior to multivariate analysis. Principal component analysis (PCA) was performed using singular value decomposition to obtain orthogonal linear combinations of the original variables that maximise explained variance. The first two principal components were used for visualisation of global variance structure. Convex hulls were applied to illustrate within-group dispersion; these were descriptive and not inferential.

Linear discriminant analysis (LDA) was subsequently performed to identify the linear combination of features that maximised separation between cancer and non-cancer groups. For two classes, LDA produces a single discriminant axis defined by maximising the ratio of between-class scatter to within-class scatter. Projection onto this axis was used to visualise supervised class separation.

Mahalanobis distances were calculated, scripted in Python, using pooled within-class covariance. Class-specific covariance matrices were computed and combined into a pooled estimate:

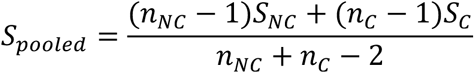

The inverse pooled covariance matrix was used to compute covariance-adjusted distances of each observation to both class centroids:

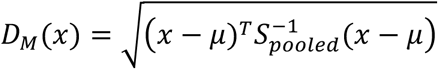

Distances were visualised as a heatmap to illustrate relative proximity to each class centroid.

### Machine Learning Analysis

To address class imbalance inherent to real-world screening cohorts, the Synthetic Minority Oversampling Technique (SMOTE; generation of synthetic minority-class samples in feature space) was applied to the training data only (10). Machine learning models were optimised using grid-search hyperparameter tuning with five-fold cross-validation, executed on a high-performance computing cluster (SCIAMA, University of Portsmouth).

Two ensemble classifiers were evaluated: a **stacked ensemble** (Random Forest and XGBoost base learners with logistic regression meta-learner) and a **soft voting ensemble** (Random Forest, XGBoost, logistic regression). Models were trained on SMOTE-balanced data and evaluated on held-out test sets. Full methodological details are provided in the online-Supplementary file.

### Metabolomic Analysis

EBC samples underwent methanol-based metabolite extraction followed by liquid chromatography–mass spectrometry (LC–MS). Data were normalised, transformed, and analysed using supervised multivariate methods to identify discriminative metabolic features. Comprehensive metabolomic workflows and analytical parameters are described in the Supplementary Methods.

## RESULTS

### Study Population and Clinical Characteristics

Staggered recruitment of participants (cases and controls) following written informed consent to the ExPeL study. The recruitment pathway is summarised in Figure 1, among lung-cancer cases, 83% had early-stage disease (Stage I–II). Mean age was similar between cases and controls (69.9 vs 68.4 years), with a slightly higher proportion of males among controls (64% males vs 48% females). Subsets used for Inflammacheck® machine-learning analysis and LC– MS metabolomics were broadly representative of the full cohort in terms of age and sex, supporting internal consistency across analyses.

**Figure 1.**
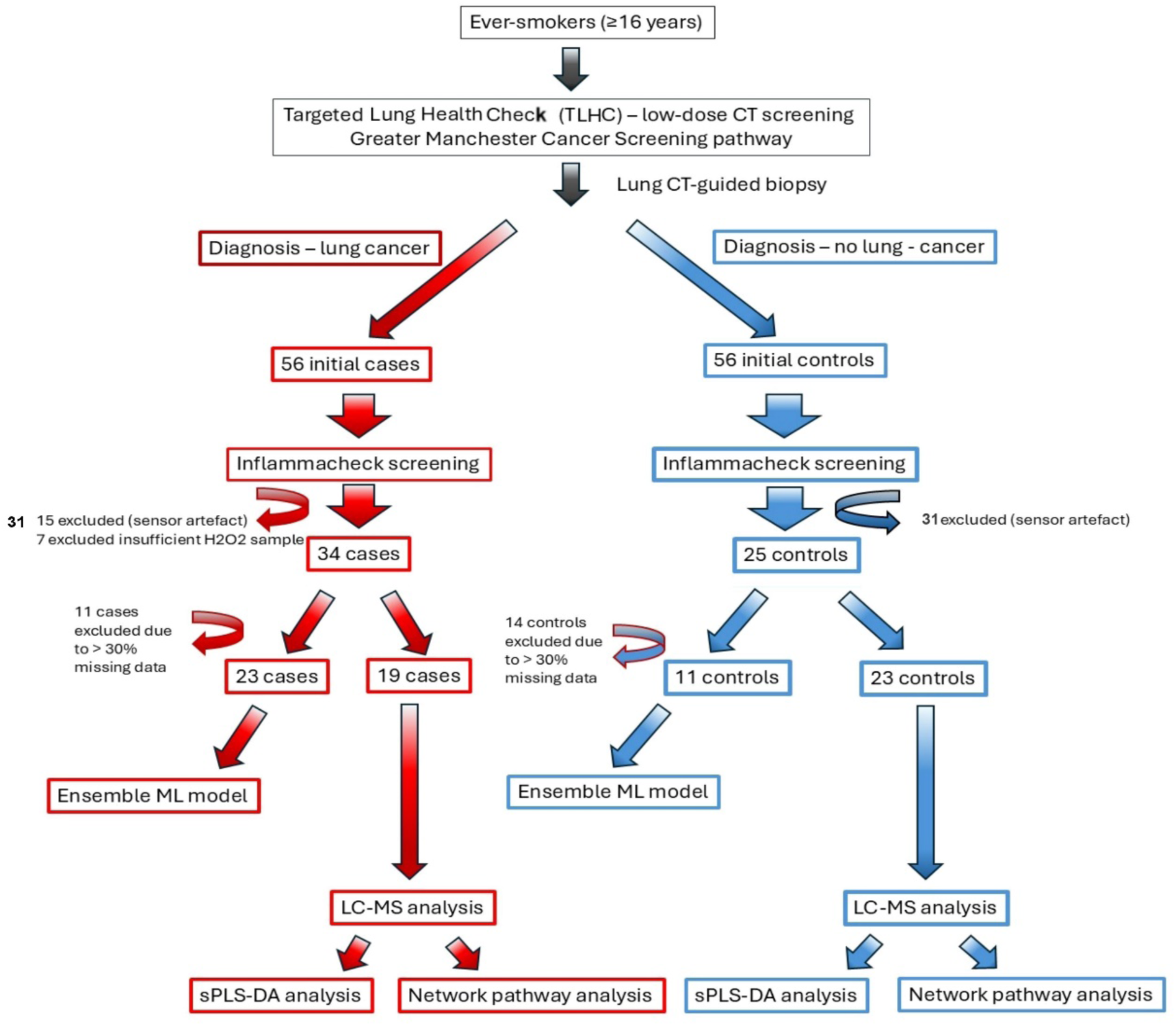
Schematic diagram showing the recruitment of participants into the ExPeL study. Participants; cases (in red) and controls (in blue), were recruited from ever-smokers who were invited to participate in the national screening for lung-cancers via the Targetted Lung Health Check programme, offering low-dose CT screening for early-detection of lung-cancers. Onward management of patients is guided primary by the NHS-England’s TLHC protocol, the British Thoracic Society (BTS) pulmonary nodule guidelines and the local lung cancer Multi-disciplinary team (MDT) processes. Patients referred for onward management of suspected lung-cancers are referred for CT-guided biopsy at secondary or tertiary referral centres. A patient is placed on the cancer-pathway for management if the histology confirms a malignancy, of there is a positive cytology sample (EBUS-sample). These participants are the cases (in red) who eventually are recruited (following written informed consent) for Inflammacheck® screening or for providing a sample to be analysed by LC-MS. Participants who have a nodule size < 5mm or < 80 mm3 are discharged from the suspected-cancer-pathway with no need for subsequent-follow-up (as per BTS guidelines). Patients from this subset were approached to participate in Inflammacheck® screening and to provide samples for LC-MS analysis following written informed consent.

Comorbidity profiles differed between cases vs control groups. Diabetes (34–39% vs 4–18%) and ischaemic heart disease (16–26% vs 0–9%) were more prevalent among cases, while prior non-lung malignancies were more common in controls (24–27%). Chronic respiratory disease prevalence (COPD and asthma) was comparable. Smoking history was similar across groups, with the majority being former smokers.

Most lung cancers were detected through screening pathways (47–52%), with the remainder identified incidentally or via symptomatic presentation. Early-stage disease predominated (58–65% Stage I), and adenocarcinoma was the most common histological subtype (39–48%), followed by squamous cell carcinoma and small cell lung cancer. These distributions were preserved across analytical subsets.

### Exploratory Multivariate Separation of Cancer and TLHC-Excluded Participants

Unsupervised principal component analysis (PCA) of standardised exhaled breath condensate (EBC) variables demonstrated partial but discernible separation between patients with confirmed lung cancer and participants excluded from further investigation within the Greater Manchester Targeted Lung Health Check (TLHC) programme (Figure 2A). Projection onto the first two principal components (PC1 and PC2), which capture the greatest proportion of overall variance in the dataset, revealed distinct geometric structuring of the two groups within multivariate space.

**Figure 2.**
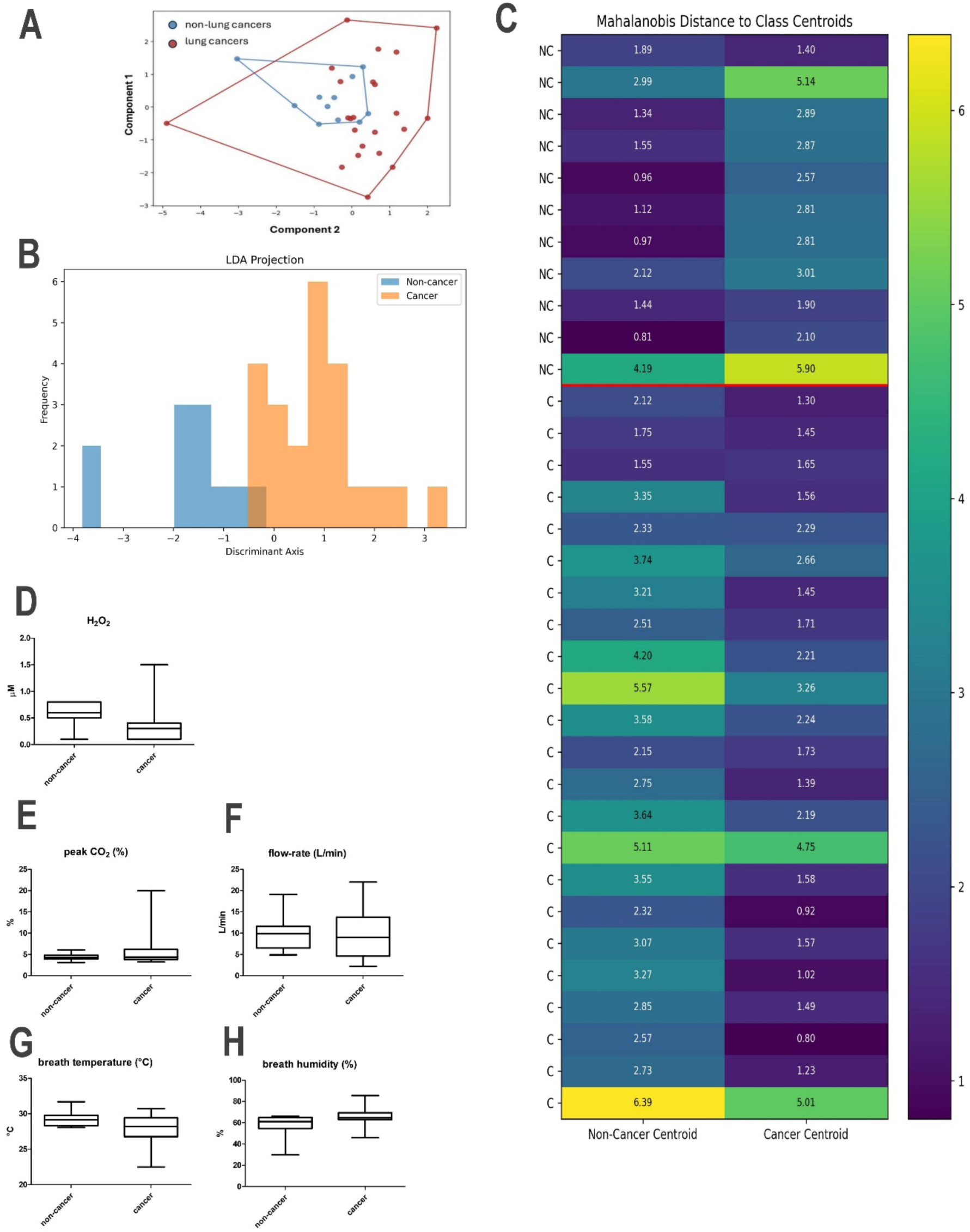
Multivariate separation of lung cancer and TLHC-excluded participants in EBC feature space and univariate comparison of exhaled breath condensate (EBC)–derived physiological and biochemical variables between cancer and non-cancer participants. (A) Principal component analysis (PCA) of standardised EBC variables showing projection onto the first two principal components. Convex hulls depict within-group dispersion. (B) Linear discriminant analysis (LDA) projection onto the single discriminant axis maximising between-class separation. (C) Heatmap of Mahalanobis distances from each participant to cancer and non-cancer class centroids using pooled covariance, illustrating covariance-adjusted multivariate proximity. (D–H) Box-and-whisker plots summarising individual EBC-derived variables for non-cancer and cancer participants: (D) hydrogen peroxide (H₂O₂), (E) peak end-tidal CO₂ (%), (F) exhalation flow rate (L/min), (G) breath temperature (°C), and (H) breath humidity (%). Boxes represent the interquartile range (IQR; 25th–75th percentiles), the central line denotes the median, and whiskers indicate the minimum and maximum observed values. Across variables, distinct distributional patterns are evident. Non-cancer participants exhibit higher median H₂O₂ concentrations with relatively constrained variability, whereas cancer cases show lower central tendency but increased dispersion, including extreme values. Peak CO₂ demonstrates a broader range and higher upper extremes in cancer participants. Flow rate is markedly more variable in cancer cases, spanning both low and high values, while non-cancer values remain comparatively restricted. Breath temperature in cancer participants is modestly lower on average and more heterogeneous, and breath humidity displays a higher median and wider spread relative to controls. Collectively, these findings highlight increased physiological and biochemical variability in cancer participants, consistent with the multivariate dispersion noted in PCA, LDA and the heatmap analysis.

Cancer cases exhibited greater dispersion, occupying a broader region of principal component space, whereas TLHC-excluded participants were comparatively more compactly clustered. The convex hulls drawn around each group illustrate this difference in within-group variance and visually demonstrate that, although some overlap exists, this is to be expected in a real-world feasibility cohort, the groups are not randomly intermixed. Importantly, PCA is an unsupervised transformation that maximises total variance without knowledge of class labels; therefore, any visible separation observed in PCA space reflects a true intrinsic (mathematically governed) multivariate difference rather than an artificial model-driven discrimination.

To formally test whether a linear axis exists that maximally separates the two groups, linear discriminant analysis (LDA) was performed (Figure 2B). Unlike PCA, LDA is a supervised projection that identifies the axis maximising the ratio of between-class variance to within-class variance. In this binary setting, LDA yields a single discriminant axis. Projection onto this axis demonstrated clear directional separation between cancer and non-cancer participants, indicating the presence of an intrinsic linear combination of EBC features along which group centroids are maximally distinct. The observed separation is therefore not merely a function of variance structure but reflects, again, an intrinsic property of the two groups with a mathematically defined discriminative axis.

To further quantify multivariate separation while accounting for covariance structure, Mahalanobis distances (11), were computed from each individual data-point to both the cancer and non-cancer class centroids (Figure 2C). Mahalanobis distance incorporates feature correlations and scaling, providing a covariance-adjusted metric of geometric proximity in high-dimensional space. The resulting heatmap demonstrates that, in the majority of cases, individuals are closer (in Mahalanobis space) to the centroid of their true class than to that of the opposing class. This pattern indicates coherent class-specific clustering and supports the presence of an intrinsic multivariate separation between the cancer and control participants in the ExPeL study. Collectively, the PCA geometry, LDA discriminant axis, and covariance-adjusted distance mapping converge on a consistent finding: cancer and TLHC-excluded participants occupy distinguishable regions of EBC feature space.

Although TLHC-excluded participants were not defined by long-term benign follow-up but rather by screening exclusion criteria under BTS and NICE guidance(12), the observed multivariate separation suggests that the EBC signal reflects intrinsic physiological differences between confirmed malignancy and screening-negative individuals. These findings support the biological plausibility of EBC-derived features as discriminative biomarkers in this feasibility cohort.

### Distribution of Individual EBC and Breath Physiology Variables

Comparisons of individual EBC-derived variables revealed differences in distributional patterns between cancer and non-cancer participants (Figure 2 B-F). Non-cancer participants showed higher median hydrogen peroxide concentrations, while cancer cases exhibited lower central tendency with greater dispersion, including a high outlier. Peak end-tidal CO₂ values displayed a wider range and higher upper extremes among cancer cases. Exhalation flow rate varied substantially in cancer participants, spanning both low and high values, whereas non-cancer values were more constrained. Breath temperature in cancer cases was modestly lower on average and more variable, and humidity showed a higher median and broader spread relative to controls.

### Performance of Ensemble Models for EBC-Based Classification

EBC features were used to train supervised machine-learning models to classify and predict cancer versus non-cancer status. The Voting Ensemble Model outperformed the Stacking Ensemble Model across multiple metrics (Table 2). The voting model achieved an accuracy of 85.7%, with perfect specificity (1.000) and precision (1.000), indicating strong discrimination of controls and minimal false positives. Its Matthews correlation coefficient (MCC) was 0.73, with a ROC–AUC of 0.90.

**Table 1.**
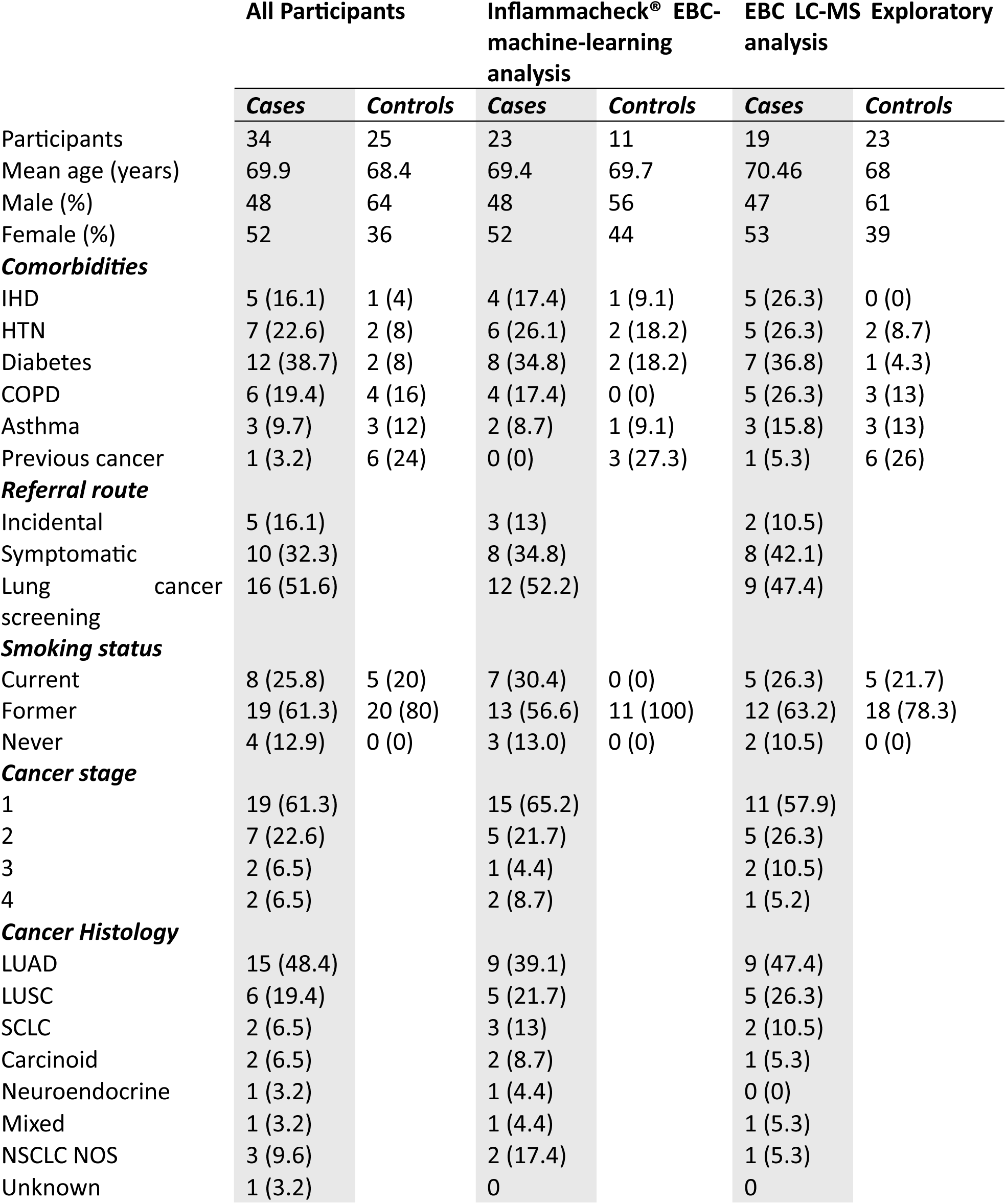
Participant demographics and clinical characteristics across overall cohort and analytical subsets. Data are presented as n (%). The table summarises age, sex, comorbidities, referral route, smoking status, lung cancer stage, and histology for all participants, as well as for the subsets used in Inflammacheck® exhaled breath condensate (EBC) machine-learning analysis and LC-MS metabolomics exploratory analysis. Comorbidities include ischemic heart disease (IHD), hypertension (HTN), diabetes, chronic obstructive pulmonary disease (COPD), asthma, and prior non-lung cancers. Lung cancer histologies include lung adenocarcinoma (LUAD), squamous cell carcinoma (LUSC), small cell lung cancer (SCLC), carcinoid, neuroendocrine, mixed, and non-small cell lung cancer not otherwise specified (NSCLC NOS).

**Table 2.** Performance Metrics for Ensemble Models Predicting Lung Health Outcomes from EBC Profiles. Comparison of the classification performance of two ensemble models — a Voting Ensemble and a Stacking Ensemble, on test data derived from SMOTED EBC features, distinguishing lung pathology (cases) vs control status. Metrics include standard binary classification indices: Accuracy, Sensitivity, Specificity, predictive values (PPV, NPV), error rates, MCC as a balanced correlation measure under class imbalance, and ROC-AUC for discriminatory ability.

| Metric | Voting Ensemble Model | Stacking Ensemble Model |
| --- | --- | --- |
| Accuracy (%) | 85.71 | 71.43 |
| Sensitivity / Recall | 0.800 | 0.800 |
| Specificity | 1.000 | 0.500 |
| Precision (PPV) | 1.000 | 0.800 |
| Negative Predictive Value (NPV) | 0.667 | 0.500 |
| False Positive Rate (FPR) | 0.000 | 0.500 |
| Matthews Correlation Coefficient (MCC) | 0.730 | 0.300 |
| ROC-AUC Score | 0.900 | 0.900 |
| Misclassification Rate (%) | 14.29 | 28.57 |
| Confusion Matrix (TN, FP; FN, TP) | (2, 0; 1, 4) | (1, 1; 1, 4) |

In contrast, the stacking ensemble achieved an accuracy of 71.4%, lower specificity (0.50), and an MCC of 0.30, while maintaining similar sensitivity and ROC–AUC. These results demonstrate superior overall performance and stability of the voting ensemble in this dataset.

### Untargeted LC–MS Metabolomic Profiling of EBC

Untargeted liquid chromatography–mass spectrometry (LC–MS) analysis of EBC identified 2,132 molecular features across samples. Using sparse partial least squares discriminant analysis (sPLS-DA), classification accuracy of 84.1% was achieved (Figure 3A). Features were ranked using variable importance in projection (VIP) scores, and the top four discriminative molecules were selected (Figure 3B–E).

**Figure 3.**
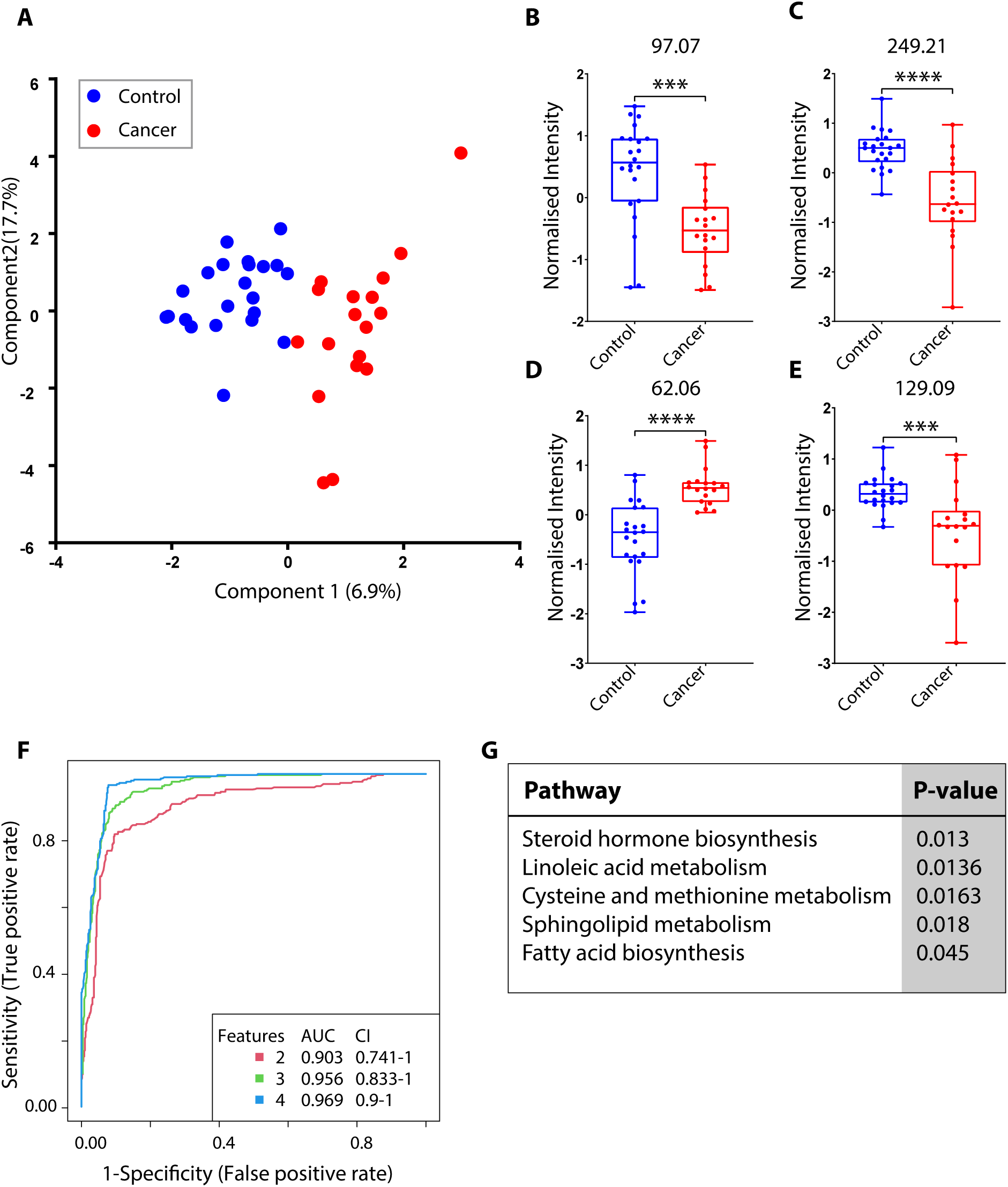
Exhaled Breath condensate exploratory analysis. A. sPLS-DA scores plot using two components and 10 features per component. Average error rate of classification was 15.9%, in classifying non-cancer controls (blue dots) against lung cancers (red dots) participants. B-E show box plots comparing means of four m/z values 97.97, 249.21, 62.06 and 129.09, which were top four features selected based on VIP scores (VIP>1) and t-test (p<0.001) combined. F Multivariate receiver operating characteristic (mROC) curve using PLS-DA and combination of 2, 3 and 4 features. The AUC was observed to be between 0.95 to 0.99 when incrementally changing from 2 to 4 features to perform the analysis. G Functional pathway analysis performed using mummichog algorithm (27), with a p-value cut-off threshold of 0.25.

Three of the four features were identified with high confidence. Tri(propylene glycol) butyl ether was annotated at Metabolomics Standards Initiative (MSI) level 3 based on spectral library matching. Two additional features were assigned molecular formulas C₇H₁₀O (putatively Hept-4-yn-2-one) and C₂H₆NO, although definitive structural annotation was not possible. These four metabolites differentiated cancer from control samples with an AUC of 0.969 (Figure 3F). Smoking status did not confound this analysis.

Network-based pathway analysis revealed enrichment of metabolic pathways including sphingolipid and fatty acid biosynthesis (Figure 3G). These metabolites represent volatile or semi-volatile compounds detectable in EBC during tidal breathing, highlighting the capacity of LC–MS profiling to capture biochemical signatures present in exhaled breath.

## DISCUSSION

The ExPeL study demonstrates that it was possible to distinguish early-stage lung cancers from at-risk controls via machine learning algorithms that were trained on exhaled breath condensate (EBC) data. EBC-analyses is a non-invasive, tidal-breathing based measurement, and for this study, we used the Inflammacheck**®** device. By integrating physiological breath features (oxidative stress, gas exchange, airflow, temperature, and humidity) with molecular profiling of volatile organic compounds (VOCs) captured during normal tidal breathing, this pilot study highlights a rapid, low-cost, safe, and scalable adjunct to low-dose CT (LDCT) screening within the UK’s Targeted Lung Health Check (TLHC) programme. The overarching aim of this study is to support early detection and prioritisation for imaging, improving efficiency in resource-intensive national screening programmes.

### Relevance of Control Definition and Real-World Screening Context

Control participants were defined as ever-smokers undergoing TLHC-based CT assessment who were not referred for further diagnostic escalation, consistent with NICE and British Thoracic Society guidance for low-risk pulmonary nodules (9). This risk-based classification provides a biologically and clinically relevant comparator, reflecting the population that would realistically be triaged in a screening context. Consequently, the differences in EBC features observed in this study correlate to early pathological variations rather than overt, advanced disease, enhancing the early-stage translational validity of our findings.

### Physiological EBC Signatures

Analysis of EBC variables revealed increased dispersion in hydrogen peroxide, end-tidal CO₂, exhalation flow, temperature, and humidity among cancer cases, consistent with clinically observed as cough, wheeze, and exertional dyspnoea, secondary to localised airway oxidative stress, reduced exercise tolerance due to inefficient ventilation–perfusion matching, and inflammatory tone in early malignancy. These subtle physiological variations are difficult to detect using conventional univariate methods, highlighting the importance of supervised machine-learning models to process and compute multivariate patterns.

When considering exhaled breath condensate H2O2 levels in early-stage, newly diagnosed cancer patients versus controls, we acknowledge the powerful influence of confounding factors. Newly diagnosed lung-cancer patients who might be clinically symptomatic following spirometry and FeNO assessments would normally be prescribed steroid inhalers. The steroid in these inhalers potentially might downregulate the production of NF-κB (13,14). By suppressing NF-κB, steroid inhalers could reduce the transcription of many inflammatory genes, leading to an overall dampening of the inflammatory response in the airways. Since H2O2 in EBC is primarily considered a marker of oxidative stress and inflammation in the airways (15,16), the anti-inflammatory effects of steroid inhalers directly lead to a reduction in EBC H2O2 levels(17).

The contrasting observations between the early-stage disease in this ExPeL study and more advanced lung-cancer cohorts such as noted in the VICTORY study (18), may reflect stage-dependent shifts in airway oxidative biology. While early malignancy may exhibit subtle or confounded oxidative signatures, advanced lung cancer is associated with a strongly pro-oxidative microenvironment in which tumour-driven reactive oxygen species production (19), outweighs any inhibitory influences, resulting in higher exhaled H₂O₂ levels (6,20).

### Machine Learning Insights

The Voting Ensemble Model, trained on SMOTE-balanced EBC data to address class imbalance in a small sample, achieved >85% accuracy, perfect specificity (1.0), high precision (1.0), and robust balanced performance (MCC 0.73, ROC-AUC 0.90). Importantly, the accuracy metric should not be misinterpreted as approximately 20% of cancers being “missed”, as these results reflect model performance within a low-prevalence, hold-out test set; in real-world TLHC populations, where cancer prevalence is low (∼1–2% per year among screened ever-smokers in Greater Manchester), predictive values must be interpreted in context. High specificity and precision are particularly valuable in screening, as they minimise unnecessary downstream imaging and reduce patient anxiety and resource burden.

### Molecular VOC Signatures from LC–MS Metabolomics

Untargeted LC–MS analysis of EBC identified over 2,000 features, from which four key VOCs were able to discriminate cancer from control samples with an AUC of 0.969. These metabolites, detected during normal tidal breathing, provide a glimpse into the biochemical microenvironment of the respiratory tract.

We have also identified individual molecules that are associated with lung cancer, which will be added to inflammacheck in the future to improve performance. Two of the top four differentially expressed molecules could be identified. Tri(propylene glycol) butyl ether is an organic solvent that is used in multiple cleaning products, meaning that most people are likely to be regularly exposed (21). It is not known to be produced as a product of intrinsic metabolism; hence it is most likely that one of the enzymes involved in its degradation pathway has been disrupted by cancer leading to over representation in the cases in this study. C7H10O was identified as Hept-4YN-2-one, for which there is little in the literature. Related molecules have been detected as part of a discriminator in the urine of patients with lung cancer previously (22).

The pathway analysis on all differentially expressed molecules provided insight into alterations in the respiratory tract in early-stage lung cancer. These included up-regulation of systems well characterised in cancer such as sphingolipids (23) and lipid metabolism (24). The other pathways identified are less established, but there are studies supporting their roles in lung cancer. For example, linoleic acid expression in platelets has previously been suggested as a biomarker of non-small cell lung cancer (25) and steroid biosynthesis has been demonstrated in lung cancer cell lines, which may have a role in immunomodulation(26).

### Clinical Implications and Future Directions

Although a proportion of TLHC-excluded participants had small or low-risk pulmonary nodules that did not meet criteria for further escalation, classification within the present study reflects their clinical status at the point of discharge under BTS-guided screening pathways. Thus, the observed discrimination between cancer and non-cancer groups pertains to contemporaneous clinical decision-making rather than retrospective confirmation of long-term benignity. In this context, the identification of intrinsic multivariate separation supports the potential utility of an adjunct physiological assay, such as Inflammacheck, to provide an additional layer of biological risk stratification alongside imaging. This may be particularly valuable not only within structured screening programmes but also in primary care settings, where general practitioners are frequently required to decide whether to refer patients presenting with non-specific yet concerning symptoms—such as persistent cough, unexplained weight loss, or haemoptysis, that often overlap with benign respiratory conditions. An accessible adjunct test capable of reflecting underlying biological perturbation could therefore help reduce diagnostic uncertainty in this grey clinical zone, supporting more informed referral decisions while avoiding unnecessary escalation in low-risk presentations.

We propose the feasibility of integrating EBC-based physiological and VOC signatures into a predictive model in future, the ExPeL study provides a proof-of-concept for a non-invasive triage tool upstream of LDCT.

Such an approach could prioritise high-risk individuals for imaging, improving the yield of positive diagnoses per scan. It may also reduce radiation exposure and healthcare resource burden, particularly in low-prevalence screening populations. The results of this study may enable mechanistically informed biomarker selection, leading to improvements of the Inflammacheck® platform.

Future work will require prospective validation in larger multi-centre trials employing TLHC populations, evaluating performance across diverse patient subgroups and exploring integration of newly identified VOCs into the predictive algorithm. By combining **machine** learning, multivariate physiological signals, and VOC metabolomics, EBC-based approaches hold promise as scalable, rapid, and clinically actionable tools to enhance early lung cancer detection.

### Limitations of the study

A key limitation of this study relates to the definition of the control group. Controls were derived from screen-negative, low-risk participants within the Greater Manchester Targeted Lung Health Check programme rather than individuals with confirmed long-term benign status. While some participants had small or low-risk pulmonary nodules classified according to established clinical guidance, discharge from the screening pathway reflects real-world decision-making in contemporary CT-based lung cancer screening programmes. Therefore, our control cohort represents a pragmatic screening population rather than a lifelong benign comparator, which should be considered when interpreting diagnostic performance estimates.

## Data Availability

All data produced in the present study are available upon reasonable request to the authors

## ACKNOWLEDGEMENTS

We thank Helle Funch Nielsen and Stig Lytke Brejl for their invaluable scientific, engineering and partnership expertise. This study was supported by Exhalation Technology Ltd through in-kind contributions of test devices and kits. We would also like to thank the Acute and Inpatient research teams at the Northern Care Alliance Foundation Trust for their hard work in delivering this study. This study was sponsored by the Northern Care Alliance Foundation Trust and funded by the Wellcome Trust.

## REFERENCES

1. Gracie K, Kennedy MPT, Esterbrook G, Smith G, Blaxill P, Ameri AT, Rodger KIA, Robson JM, Paramasivam E, Naseer R, et al. The proportion of lung cancer patients attending UK lung cancer clinics who would have been eligible for low-dose CT screening. Eur Respir J (2019) 54: doi: 10.1183/13993003.02221-2018

2. Peralbo-Molina A, Calderón-Santiago M, Priego-Capote F, Jurado-Gámez B, Luque De Castro MD. Metabolomics analysis of exhaled breath condensate for discrimination between lung cancer patients and risk factor individuals. J Breath Res (2016) 10:016011. doi: 10.1088/1752-7155/10/1/016011

3. Peralbo-Molina A, Calderón-Santiago M, Priego-Capote F, Jurado-Gámez B, Luque De Castro MD. Identification of metabolomics panels for potential lung cancer screening by analysis of exhaled breath condensate. J Breath Res (2016) 10:026002. doi: 10.1088/1752-7155/10/2/026002

4. Weinberg F, Ramnath N, Nagrath D. Reactive Oxygen Species in the Tumor Microenvironment: An Overview. Cancers 2019, Vol 11, (2019) 11: doi: 10.3390/CANCERS11081191

5. Chan HP, Tran V, Lewis C, Thomas PS. Elevated levels of oxidative stress markers in exhaled breath condensate. Journal of Thoracic Oncology (2009) 4:172–178. doi: 10.1097/JTO.0b013e3181949eb9

6. Kolbasina NA, Gureev AP, Serzhantova O V., Mikhailov AA, Moshurov IP, Starkov AA, Popov VN. Lung cancer increases H2O2 concentration in the exhaled breath condensate, extent of mtDNA damage, and mtDNA copy number in buccal mucosa. Heliyon (2020) 6: doi: 10.1016/J.HELIYON.2020.E04303

7. Lee W, Thomas PS. Oxidative Stress in COPD and its measurement through exhaled breath condensate. Clin Transl Sci (2009) 2:150–155. doi: 10.1111/J.1752-8062.2009.00093.X;SUBPAGE:STRING:ABSTRACT;REQUESTEDJOURNAL:JOURNAL:17528062;JOURNAL:JOURNAL:17528062;WGROUP:STRING:PUBLICATION

8. Fox L, D’Cruz LG, Chauhan M, Gates J, Szarazova N, De Vos R, Hicks A, Brown T, Stores R, Chauhan AJ. Diagnosis of respiratory conditions using exhaled breath condensate using Inflammacheck® and advanced analytics: insights from the VICTORY study. J Breath Res (2025) 19:036005. doi: 10.1088/1752-7163/ADD17C

9. NHS England » Standard protocol and quality assurance standards for the Lung Cancer Screening Programme. https://www.england.nhs.uk/publication/targeted-screening-for-lung-cancer/?utm_source=chatgpt.com [Accessed January 26, 2026]

10. Chawla N V., Bowyer KW, Hall LO, Kegelmeyer WP. SMOTE: Synthetic Minority Over-sampling Technique. Journal of Artificial Intelligence Research (2002) 16:321–357. doi: 10.1613/JAIR.953

11. Mahalanobis PC. ON THE GENERALIZED DISTANCE IN STATISTICS on JSTOR. Proc Natl Inst Sci India (1936) 2:49–55. https://www.jstor.org/stable/48723335 [Accessed March 4, 2026]

12. Callister MEJ, Baldwin DR, Akram AR, Barnard S, Cane P, Draffan J, Franks K, Gleeson F, Graham R, Malhotra P, et al. British Thoracic Society guidelines for the investigation and management of pulmonary nodules: accredited by NICE. Thorax (2015) 70:ii1–ii54. doi: 10.1136/THORAXJNL-2015-207168

13. Hancox RJ, Stevens DA, Adcock IM, Barnes PJ, Taylor DR. Effects of inhaled beta agonist and corticosteroid treatment on nuclear transcription factors in bronchial mucosa in asthma. Thorax (1999) 54:488–492. doi: 10.1136/thx.54.6.488

14. Wilson SJ, Wallin A, Della-Cioppa G, Sandström T, Holgate ST. Effects of budesonide and formoterol on NF-kappaB, adhesion molecules, and cytokines in asthma. Am J Respir Crit Care Med (2001) 164:1047–1052. doi: 10.1164/ajrccm.164.6.2010045

15. Emelyanov A, Fedoseev G, Abulimity A, Rudinski K, Fedoulov A, Karabanov A, Barnes PJ. Elevated concentrations of exhaled hydrogen peroxide in asthmatic patients. Chest (2001) 120:1136–1139. doi: 10.1378/chest.120.4.1136

16. Dohlman AW, Black HR, Royall JA. Expired breath hydrogen peroxide is a marker of acute airway inflammation in pediatric patients with asthma. Am Rev Respir Dis (1993) 148:955–960. doi: 10.1164/ajrccm/148.4_Pt_1.955

17. Oliveira-Marques V, Marinho HS, Cyrne L, Antunes F. Role of hydrogen peroxide in NF- kappaB activation: from inducer to modulator. Antioxid Redox Signal (2009) 11:2223–2243. doi: 10.1089/ars.2009.2601

18. Fox L, D’Cruz LG, Chauhan M, Gates J, Szarazova N, De Vos R, Hicks A, Brown T, Stores R, Chauhan AJ. Diagnosis of respiratory conditions using exhaled breath condensate using Inflammacheck® and advanced analytics: insights from the VICTORY study. J Breath Res (2025) 19: doi: 10.1088/1752-7163/ADD17C,

19. Papavassiliou KA, Sofianidi AA, Papavassiliou AG. Reactive Oxygen Species and the Lung Cancer Tumor Microenvironment: Emerging Therapeutic Opportunities. Antioxidants (Basel) (2025) 14: doi: 10.3390/antiox14080964

20. Kolbasina NA, Gureev AP, Serzhantova O V., Mikhailov AA, Moshurov IP, Starkov AA, Popov VN. Lung cancer increases H2O2 concentration in the exhaled breath condensate, extent of mtDNA damage, and mtDNA copy number in buccal mucosa. Heliyon (2020) 6:e04303. doi: 10.1016/J.HELIYON.2020.E04303

21. Tripropylene glycol n-butyl ether | C13H28O4 | CID 60196418 - PubChem. https://pubchem.ncbi.nlm.nih.gov/compound/1-_2-_1-butoxypropoxy_propoxy_propan-1-ol [Accessed January 26, 2026]

22. Gasparri R, Capuano R, Guaglio A, Caminiti V, Canini F, Catini A, Sedda G, Paolesse R, Di Natale C, Spaggiari L. Volatolomic urinary profile analysis for diagnosis of the early stage of lung cancer. J Breath Res (2022) 16:046008. doi: 10.1088/1752-7163/AC88EC

23. Lin M, Li Y, Wang S, Cao B, Li C, Li G. Sphingolipid Metabolism and Signaling in Lung Cancer: A Potential Therapeutic Target. J Oncol (2022) 2022:9099612. doi: 10.1155/2022/9099612

24. Eltayeb K, La Monica S, Tiseo M, Alfieri R, Fumarola C. Reprogramming of Lipid Metabolism in Lung Cancer: An Overview with Focus on EGFR-Mutated Non-Small Cell Lung Cancer. Cells 2022, Vol 11, (2022) 11: doi: 10.3390/CELLS11030413

25. de Castro J, Rodríguez MC, Martínez-Zorzano VS, Llanillo M, Sánchez-Yagüe J. Platelet linoleic acid is a potential biomarker of advanced non-small cell lung cancer. Exp Mol Pathol (2009) 87:226–233. doi: 10.1016/j.yexmp.2009.08.002

26. Merk VM, Grob L, Fleischmann A, Brunner T. Human lung carcinomas synthesize immunoregulatory glucocorticoids. Genes & Immunity 2023 24:1 (2023) 24:52–56. doi: 10.1038/s41435-023-00194-y

27. Li S, Park Y, Duraisingham S, Strobel FH, Khan N, Soltow QA, Jones DP, Pulendran B. Predicting Network Activity from High Throughput Metabolomics. PLoS Comput Biol (2013) 9:e1003123. doi: 10.1371/JOURNAL.PCBI.1003123

