## Supplementary information for "Harnessing exhaled breath for lung cancer early detection – results from the ExPeL study"

#### Supplementary Information – Detailed Definition of Control Cohort

##### Rationale for Control Classification

In this study, control participants were defined within the context of a nationally implemented lung cancer screening programme rather than by absolute absence of pulmonary abnormalities. All controls were ever-smokers assessed through the Greater Manchester TLHC pathway and underwent low-dose CT imaging as part of routine risk-stratified screening.

Individuals were classified as controls if they were not diagnosed with lung cancer following initial CT assessment and guideline-directed follow-up, and were not referred for further diagnostic investigation or continued surveillance. Eight participants had pulmonary nodules categorised as **low risk** according to established radiological features, including small size, benign morphology, or low calculated malignancy probability, consistent with national pulmonary nodule management guidance. One additional participant underwent CT-guided biopsy, which confirmed the absence of malignancy.

##### Alignment with National Guidelines

This classification approach reflects current **British Thoracic Society (BTS)** and **NICE** pulmonary nodule management recommendations, which support discharge from follow-up for nodules assessed as low risk without mandatory longitudinal imaging. As such, inclusion as a control does not imply permanent benign status but represents a **negative outcome at first-pass screening**, consistent with the clinical decision-making framework embedded within TLHC.

##### Relevance to Screening Tool Evaluation

Within this framework, exhaled breath condensate (EBC) measurements from controls capture biomarker profiles associated with nodules deemed non-malignant at screening and excluded from diagnostic escalation. This provides the biologically and clinically relevant comparator group for assessing EBC as a **rapid, non-invasive, and cost-effective adjunct to CT-based screening**, aimed at identifying individuals who warrant further investigation rather than replacing established surveillance pathways.

##### Exclusion Criteria

Exclusion criteria for both cases and controls included a diagnosis of malignancy at another site within the preceding two years, prior lung cancer treatment, acute respiratory infection at the time of sampling, or inability to provide informed consent.

### Additional information to the methodology in this study

##### Detailed Definition of Control Participants

Controls were defined within the operational framework of the TLHC programme rather than by absolute absence of pulmonary abnormalities. All control participants were ever-smokers assessed by low-dose CT imaging and classified as not requiring diagnostic escalation following programme-level radiological review. Eight participants had pulmonary nodules deemed low risk according to national guidance (e.g. small size, benign morphology, or low malignancy probability), and one participant underwent CT-guided biopsy confirming absence of malignancy.

This risk-based classification aligns with British Thoracic Society and NICE pulmonary nodule management guidelines, which permit discharge without prolonged surveillance for low-risk nodules. Accordingly, control status represents a negative screening outcome rather than long-term benign certainty.

##### Inflammacheck® and Coronacheck® Systems

The Inflammacheck® device has been previously described and measures real-time EBC-derived biomarkers reflective of airway oxidative stress and physiology. Coronacheck® is an analogous system designed to facilitate larger-volume EBC collection for metabolomic profiling.

##### Machine Learning Pipeline

EBC data were imported into Python and pre-processed using pandas and NumPy. Features were standardised using scikit-learn's StandardScaler. Stratified train–test splitting was employed to avoid information leakage. SMOTE was applied exclusively to the training set to mitigate class imbalance without inflating test performance.

Hyperparameter optimisation was conducted via exhaustive grid-search with five-fold cross-validation. Computation was accelerated using the SCIAMA supercomputing cluster (University of Portsmouth).

##### Ensemble Modelling Strategy

Stacking and voting ensemble methods were selected due to their ability to combine complementary model strengths and reduce bias and variance. The stacked model used Random Forest and XGBoost classifiers as base learners and logistic regression as a meta-learner. The voting ensemble aggregated class probabilities across Random Forest, XGBoost, and logistic regression models.

##### Metabolomic Processing and Analysis

EBC metabolites were extracted using methanol and analysed by LC–MS. Compound identification employed GNPS spectral matching and SIRIUS-based in silico fragmentation. All annotations were classified as Metabolomics Standards Initiative (MSI) level 3. Data were normalised to sample weight, log-transformed, Pareto-scaled, and analysed using sparse partial least squares discriminant analysis (sPLS-DA) in MetaboAnalyst 5.0. Feature importance was assessed using VIP scores and statistical testing, followed by pathway enrichment analysis.
